# Systematic reviews in minutes to hours using artificial intelligence

**DOI:** 10.64898/2026.02.06.26345764

**Authors:** Liam Bakker, Thomas Caganek, Amrit Rooprai, Samuel Hume

## Abstract

Systematic reviews are used in academia, biotechnology, pharmaceutical companies and government to synthesise and appraise large numbers of publications. The current (largely manual) workflow takes an average of 9-18 months^1^, at a cost of $100,000+ per review^2^. We built a platform, ScholaraAI, that leverages artificial intelligence to cut this to < 0.1% of the time, without compromising quality. ScholaraAI facilitates end-to-end systematic reviews; search, screening, data extraction, and analysis. The workflow is transparent, and the researcher is in the loop. Our approach is compliant with the PRISMA and RAISE frameworks. Compared to a benchmarking set of published systematic reviews, ScholaraAI’s sensitivity for correctly included studies is 100% ± 0%, its specificity for correctly excluded studies is 90.8 ± 8.6%, and its accuracy for data extraction is 98.0 ± 3.5%. The time taken per review was 3.67 hours ± 1.26. We used ScholaraAI to produce a novel, up-to-date systematic review and meta-analysis, which is presented here. ScholaraAI is free to try at app.scholara.ai.

## Introduction

Systematic reviews are considered the highest quality of evidence^3^. The PRISMA (Preferred Reporting Items for Systematic reviews and Meta-Analyses) guidelines mandate pre-registration of a protocol, a clear research question with explicit PICO components (Population, Intervention, Comparator, Outcome), a comprehensive search strategy, defined inclusion and exclusion criteria for study screening (usually with >1 author), extraction of pre-defined data, and pre-defined synthesis methods^4^.

Systematic reviews, which include rapid reviews, scoping reviews, and overviews, form the foundation of clinical guidelines. Beyond this, they are used by academics for research, by pharmaceutical companies for regulatory submissions, and by governments for evidence synthesis during crises.

>30,000 systematic reviews are published every year^5^, for a total time cost of 9,000,000 person-hours and an estimated $4B^1^. As well as time and economic costs, the manual nature of systematic reviews means they are prone to errors – particularly on study inclusion and data extraction^6,7^. Because they can take >1 year to complete, systematic reviews are often out-of-date as soon as they’re published.

To help solve these problems, we built ScholaraAI, to produce publication-quality PRISMA^4^ and RAISE (Responsible use of AI in evidence SynthEsis)^8^-compliant systematic reviews, with human-level quality, in < 0.1% of the time.

## Methods

### Search

Our approach follows the same steps as a classical PRISMA-compliant systematic review. Throughout, we use the foundation models from OpenAI, Anthropic and DeepMind. The user starts with their own question – for example:

**Figure 1:**
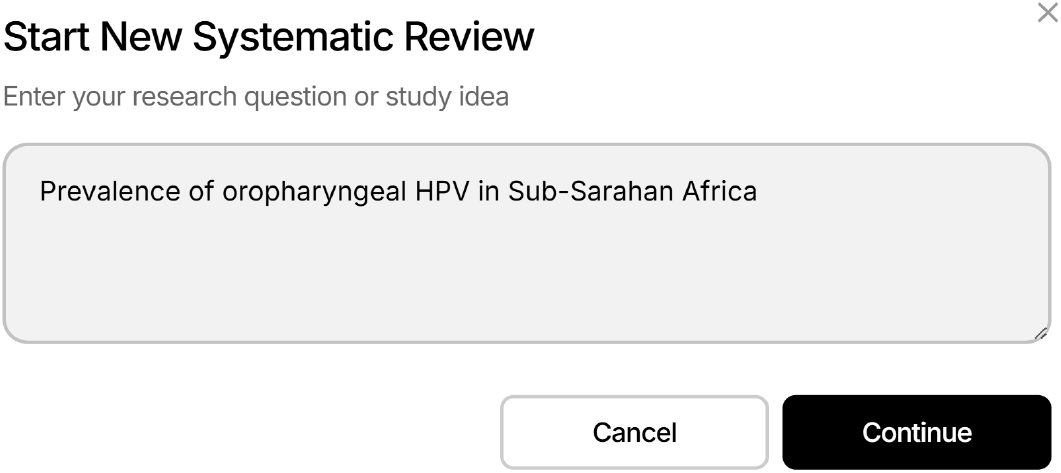
User question.

The user provides inclusion and exclusion criteria, according to the PICO framework. Claude Sonnet 4 is used to convert the user question into an editable search string, for the PubMed API:

**Figure 2:**
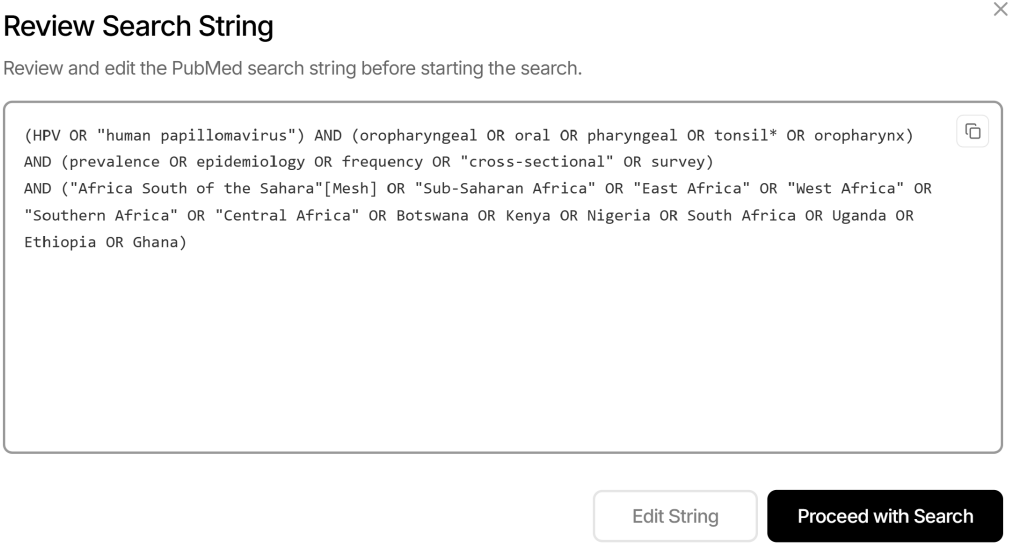
The user question is converted to a search string.

### Screening

Current systematic reviews require three humans for screening: two to make independent decisions, and a third to act as tiebreaker. Emulating this approach, three models (GPT-4o, Gemini 2.0 Flash, and Claude Sonnet 4) are used to determine study inclusion by screening titles, abstracts and full texts on the pre-defined criteria. If the models disagree, a majority vote determines inclusion (or the researcher can adjudicate). The reasons for exclusion are stated transparently:

**Figure 3:**
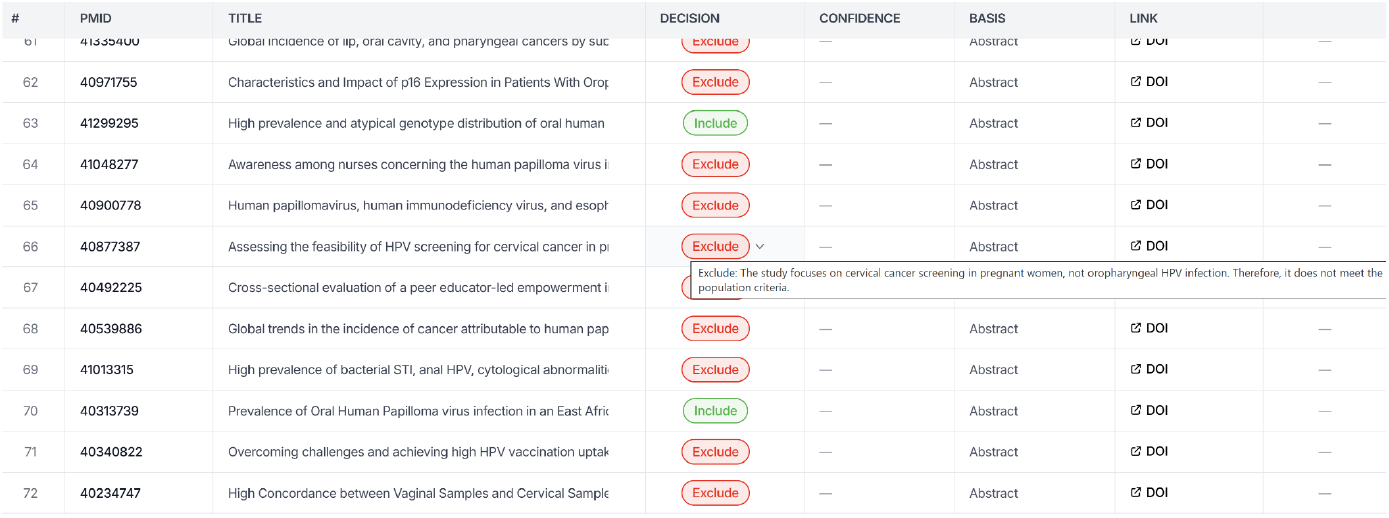
Three agents are used for study screening.

After full text screening, the user can generate a PRISMA 2020 Flow Diagram:

**Figure 4:**
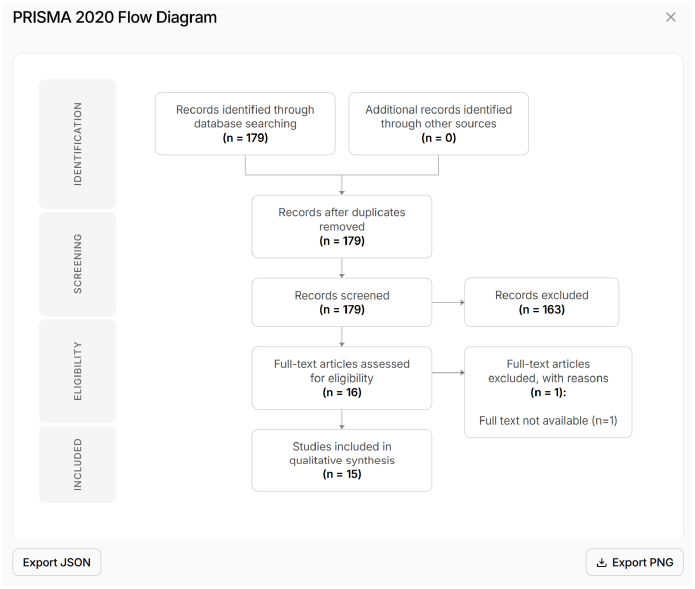
PRISMA Flow diagram.

### Data extraction and analysis

For data extraction, GPT-5.2 Vision scans PDFs, extracting numerical values with context (type, treatment arm, timepoint, source location) into a pre-built index. GPT-4o-mini matches extraction questions to the pre-scanned index. GPT-5.2 with JSON mode extracts values directly from PDFs when smart matching fails. To maximise transparency, the system highlights where in the text or tables the data have been extracted from.

Meta-analyses are conducted using a DerSimonian–Laird random-effects model, supporting risk ratios (RR), odds ratios (OR), mean differences (MD), and standardized mean differences (SMD; Hedges’ g with bias correction), with heterogeneity assessed using I^2^, tau-squared, and Cochran’s Q; forest plots were generated programmatically.

## Results

### Time taken

The time taken for a systematic review according to the current standard is ∼11 thousand hours^1^. Consistent with this, the estimated time taken for the gold standard, published systematic reviews in our benchmarking dataset^9–11^ was ∼14 thousand hours. By contrast, the average time taken to replicate the same systematic reviews using ScholaraAI was 3.67 hours ± 1.26 (Figure 5).

**Figure 5:**
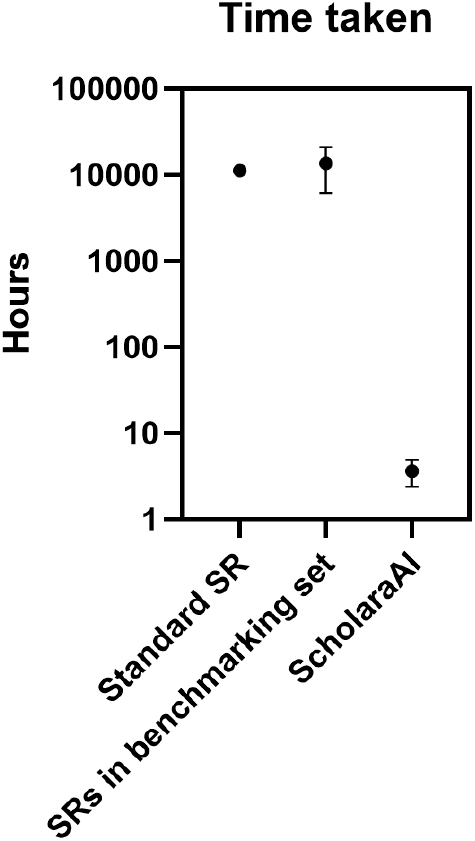
Time taken for a standard systematic review^1^, for the systematic reviews in the benchmarking set, and for the same systematic reviews on ScholaraAI. Data shown are mean ± SD.

### Sensitivity, specificity, and accuracy of data extraction

The three critical numbers in systematic review accuracy are sensitivity, specificity, and accuracy (of data extraction). These are defined as follows. Sensitivity: proportion of studies included in the ground truth study that are also included by ScholaraAI. Specificity: proportion of studies excluded in the ground truth study that are also excluded by ScholaraAI. Accuracy of data extraction: numbers must match exactly to score a point. ScholaraAI’s sensitivity was 100.0 ± 0.0%, specificity 90.8 ± 8.6%, and data extraction accuracy 98.0 ± 3.5% (Figure 6).

**Figure 6:**
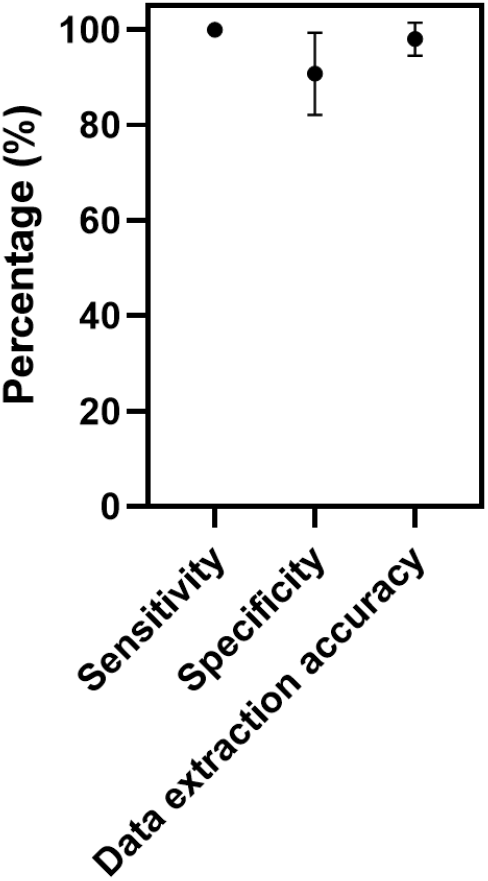
Sensitivity, specificity, and data extraction accuracy of ScholaraAI vs. a gold-standard group of published systematic reviews.

### Case study

We reproduced the study “One-Year Efficacy of Guselkumab Versus Advanced Therapies for the Treatment of Moderately to Severely Active Crohn’s Disease: A Network Meta-Analysis”, which was published in June 2025^11^. The initial search was done in July, 2023. Our search was done in January 2026.

#### Our search strategy

(“Crohn Disease”[Mesh] OR “Crohn*”[tiab]) AND (guselkumab OR infliximab OR adalimumab OR certolizumab OR vedolizumab OR ustekinumab OR risankizumab OR mirikizumab OR upadacitnib OR tofacitinib OR ozanimod OR etrasimod) AND (randomized controlled trial[pt] OR randomized[tiab] OR placebo[tiab])

#### Our inclusion criteria

Population: Adults with moderately to severely active Crohn’s disease

Intervention: Advanced therapies (guselkumab, infliximab, adalimumab, certolizumab, vedolizumab, ustekinumab, risankizumab, mirikizumab, upadacitnib, tofacitinib, ozanimod, etrasimod)

Comparator: active control or placebo

Outcomes: Clinical response

#### Our exclusion criteria

Study types: Case series; cross-sectional studies; systematic reviews; meta-analyses; qualitative studies; case reports

Participants: In vitro studies; ulcerative colitis Interventions: Protocol only

We found 285 studies in the initial search, 14 of which were ultimately included (Figure 7).

**Figure 7:**
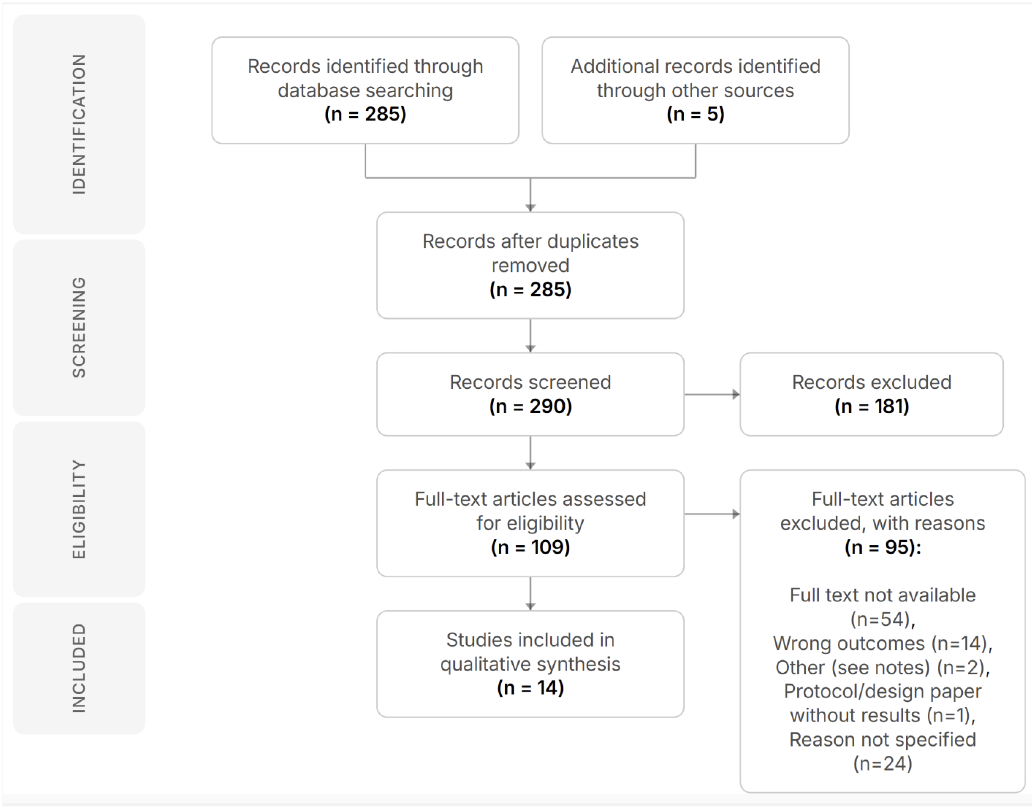
PRISMA flow chart.

In our replication, ScholaraAI included all of the studies in the initial study, as well as two studies that had since been published (the original review used data on file or conference data) and one study on a newer drug, Filgotinib^12^, that the initial study did not include (Figure 8).

**Figure 8:**
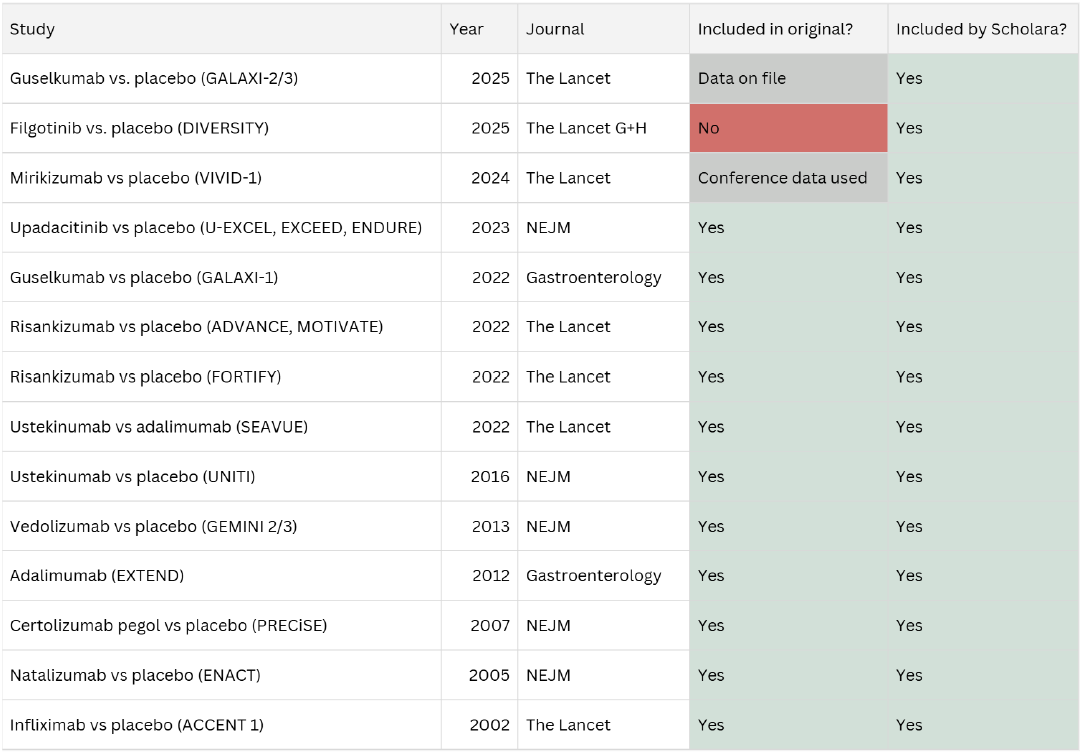
Included studies vs. the original.

We meta-analysed these data, to lay out the most to least effective biologics to induce endoscopic remission at one year in moderate to severely active Crohn’s disease (Figure 9).

**Figure 9:**
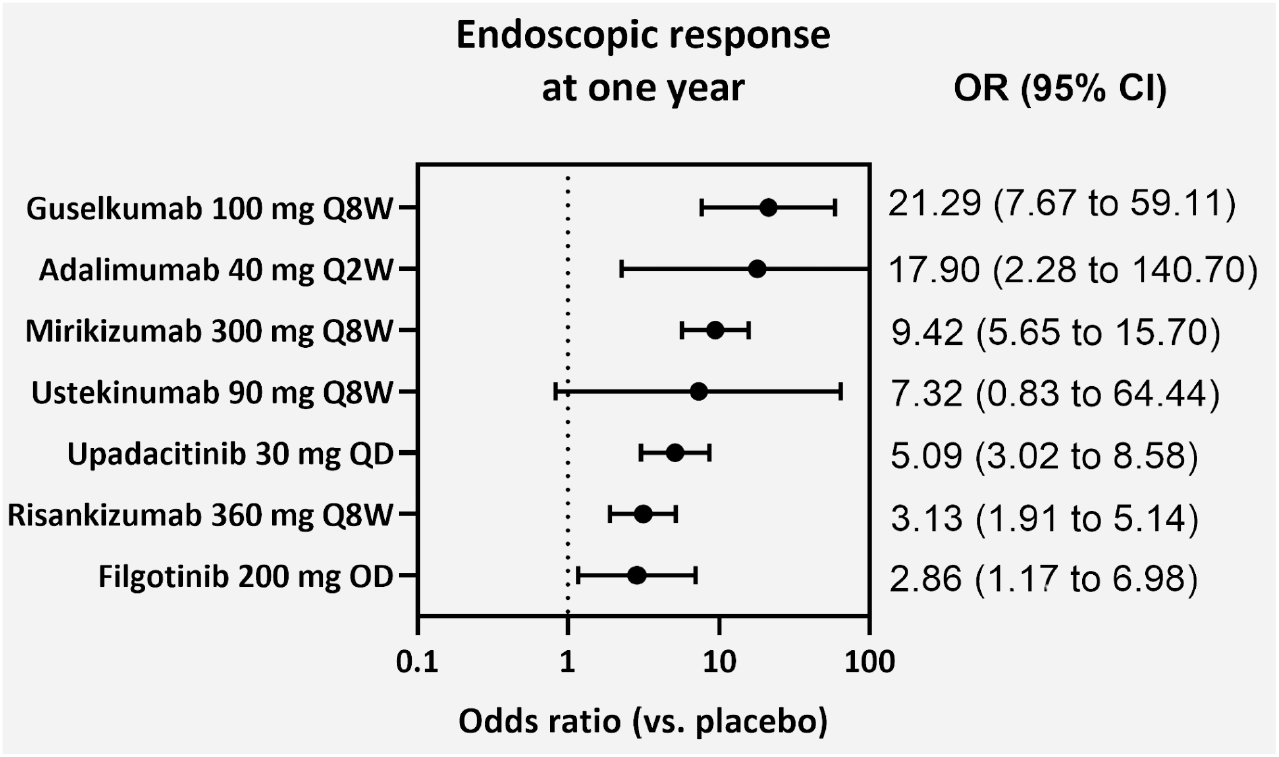
Meta-analysis for endoscopic response vs. placebo.

## Discussion

ScholaraAI leverages AI to produce PRISMA-compliant systematic reviews. It is free to try at app.scholara.ai. In our benchmarking experiments against a published set of systematic reviews, ScholaraAI has an average time of 3.67 hours ± 1.26. Its sensitivity, specificity and specificity was 100.0 ± 0.0%, 90.8 ± 8.6%, and 98.0 ± 3.5%, respectively. A feature of our model is the researcher-in-the-loop structure: reasoning is auditable and overridable.

Our approach is not without limitations. First, we only integrated ScholaraAI with a single database. Second, our benchmarking data are limited by the accuracy of the initial, ground truth data – we noted, for example, that specificity reduces for older studies, because we include more recent studies that are not in the original search. Third, we focussed only on studies in English, and the software is only available in English.

There are three particularly exciting potential use cases of our tool. First, to speed up evidence synthesis during crises (the use of systematic reviews during the Covid crisis, for example, was notoriously poor^13^). Second, to democratise access to high quality evidence synthesis globally: our tool could lower barriers for high quality evidence synthesis in low-resource institutions. Third, for *evergreen* systematic reviews that are continuously updated.

## Data Availability

All data produced in the present work are contained in the manuscript

https://app.scholara.ai/

## Ethics approval

Not required

## Software availability

app.scholara.ai

